# Ozone Generation from a Germicidal Ultraviolet Lamp with Peak Emission at 222 nm

**DOI:** 10.1101/2023.05.17.23290115

**Authors:** Michael F. Link, Andrew Shore, Behrang H. Hamadani, Dustin Poppendieck

## Abstract

Recent interest in commercial devices containing germicidal ultraviolet lamps with a peak emission wavelength at 222 nm (GUV222) has focused on mitigating virus transmission indoors and disinfecting indoor spaces while posing minimum risk to human tissue. However, 222 nm light can produce ozone (O_3_) in air. O_3_ is an undesirable component of indoor air because of health impacts from acute to chronic exposure and its ability to degrade indoor air quality through oxidation chemistry. We measured the total irradiance of one GUV222 lamp at a distance of 5 cm away from the source to be 27.0 W m^-2^ ± 4.6 W m^-2^ in the spectral range of 210 nm to 230 nm, with peak emission centered at 222 nm and evaluated the potential for the lamp to generate O_3_ in a 31.5 m^3^ stainless steel chamber. In seven four-hour experiments average O_3_ mixing ratios increased from levels near the detection limit of the instrument to 48 ppb_v_ ± 1 ppb_v_ (94 μg m^-3^ ± 2 μg m^-3^). We determined an average constant O_3_ generation rate for this lamp to be 1.10 mg h^-1^ ± 0.15 mg h^-1^. Using a radiometric method and chemical actinometry, we estimate effective lamp fluences that allow prediction of O_3_ generation by the GUV222 lamp, at best, within 10 % of the measured mixing ratios. Because O_3_ can react with gases and surfaces indoors leading to the formation of other potential by-products, future studies should evaluate the production of O_3_ from GUV222 air cleaning devices.

**TOC:** 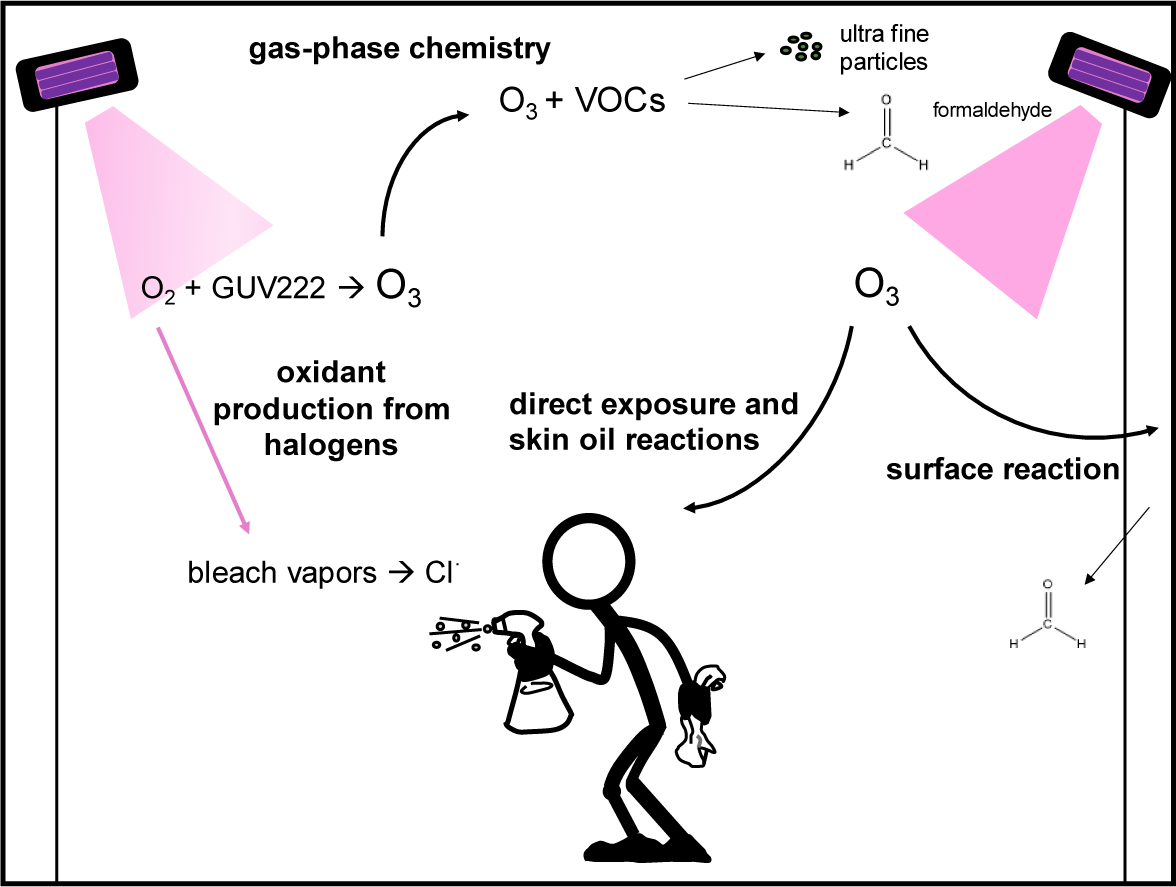

## Introduction

The on-going COVID-19 pandemic has highlighted the need for effective, in room, low energy air cleaning devices to enable safer in-person interactions in indoor environments.^1,2,3^ Portable cleaning devices use a range of technologies that may have uncharacterized impacts on indoor air quality.^4^ These impacts could result in human exposure to pollutants that are at odds with the intended benefit of the technology.^5^ One such technology is germicidal ultraviolet lamps that operate with a peak emission wavelength at 222 nm (GUV222). This wavelength is appealing as research to date indicates it does not significantly penetrate human skin and is effective at inactivating pathogens.^6,7,8^

Air cleaning devices equipped with GUV222 lamps are of particular importance when considering the potential for ozone (O_3_) formation. In the range of 175 nm to 242 nm, molecular oxygen (O_2_) will absorb light and dissociate with a quantum yield of unity to produce two ground state oxygen atoms (O), via reaction 1, that can then go on to recombine with O_2_, in a termolecular reaction involving a collisional body (M = N_2_ or O_2_), to form O_3_ via reaction 2.^9,10,11^

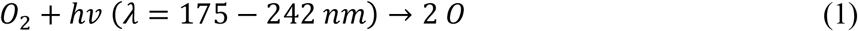

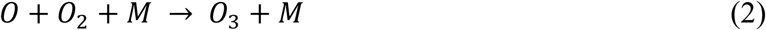

Reactions 1 and 2 are two of the four reactions comprising The Chapman mechanism which describes the production of O_3_ in the stratosphere.^12^ Characterization of spectral output and potential for O_3_ production is necessary when considering the application of GUV222 devices for mitigating virus transmission while maintaining good air quality in indoor environments.

While O_3_ itself can be a harmful by-product of air cleaner operation^13^, it can also react with gases and surfaces indoors^14^—including human skin^15^—leading to the formation of other potentially concerning by-products such as gas-phase aldehydes and ultra-fine particulate matter.^16^ Of particular concern is the exposure to O_3_ and O_3_-generated indoor pollutant by-products from application of multiple GUV222 units in small and/or poorly ventilated indoor spaces.^17^ Here we present measurements of O_3_ generation from a commercial GUV222 lamp in a stainless-steel laboratory chamber, support our O_3_ formation observations with a chemical kinetic model, and determine O_3_ generation rates for this GUV222 lamp that can be used in future evaluations of GUV222 technologies in indoor spaces.

## Methods

### Measurement of the GUV222 Lamp Emission Spectrum

Spectral irradiance measurements of a krypton chloride (KrCl) excimer GUV222 lamp were performed with a commercial UV spectrometer (Mightex Systems model: HRS-UV1-025) with detection sensitivity in the spectral range of 200 nm to 415 nm. GUV222 emission light was collected by an integrating sphere detector that was connected to the spectrometer by a UV-transmitting optical fiber patch cable. Wavelength calibration of the spectrometer was achieved by use of spectral calibration lamps with well-defined emission peaks. For the spectral irradiance calibration, an internal FEL lamp setup was used to establish the absolute scale in the 300 nm to 400 nm range, and this scale was tied (tie point at 310 nm) to an unscaled spectral calibration factor that was obtained from a deuterium lamp in the 200 nm to 340 nm range. This process yielded a continuous absolute spectral calibration factor from 210 nm to 415 nm for the UV spectrometer. We estimate the uncertainty (k=2) of the spectral irradiance measurements at 222 nm to be 17 %.

### Operation of Chamber and Experiment Design

We operated the commercial GUV222 lamp in a 31.5 m^3^ environmentally controlled walk-in chamber instrumented to measure O_3_ (Thermo 49iq O_3_ monitor) and sulfur hexafluoride (SF_6_; proton-transfer mass spectrometry) to measure the chamber air change rate (Figure S8). The O_3_ monitor was calibrated to the NIST Standard Reference Photometer prior to the study.^18^ A series of seven experiments were conducted to measure O_3_ production from the GUV222 lamp. Prior to the experimental series the chamber was passivated with 100 ppb_v_ of O_3_ for ten hours. A metal fan was placed in the chamber to facilitate mixing. The GUV222 lamp was positioned in the upper corner of the chamber pointed down and towards the center of the chamber opposite of the fan (Figure S4).

Prior to each experiment we operated the chamber to achieve a temperature of 20 °C and 50 % relative humidity. At the beginning of each experiment temperature and humidity control was stopped and the vents controlling the recirculation of air were closed. The average temperature during the experiments was 22.5 °C ± 1.3 °C, and the average relative humidity was 42.8 % ± 6.0 %. The GUV222 lamp was then turned on for four hours over which O_3_ concentration was measured. SF_6_ was injected into the chamber at the start of each experiment and air change was determined from the first order loss constant (Figure S8). Tetrachloroethylene was vaporized and introduced to the chamber at the beginning of four of the experiments to measure the effective photon flux via actinometry (e.g., Peng, et al. 2023)^19^.

## Results

### GUV222 Lamp Emission Spectrum

Figure 1a shows the spectral irradiance versus wavelength of the GUV222 lamp measured directly under and at several distances from the lamp.

**Figure 1.**
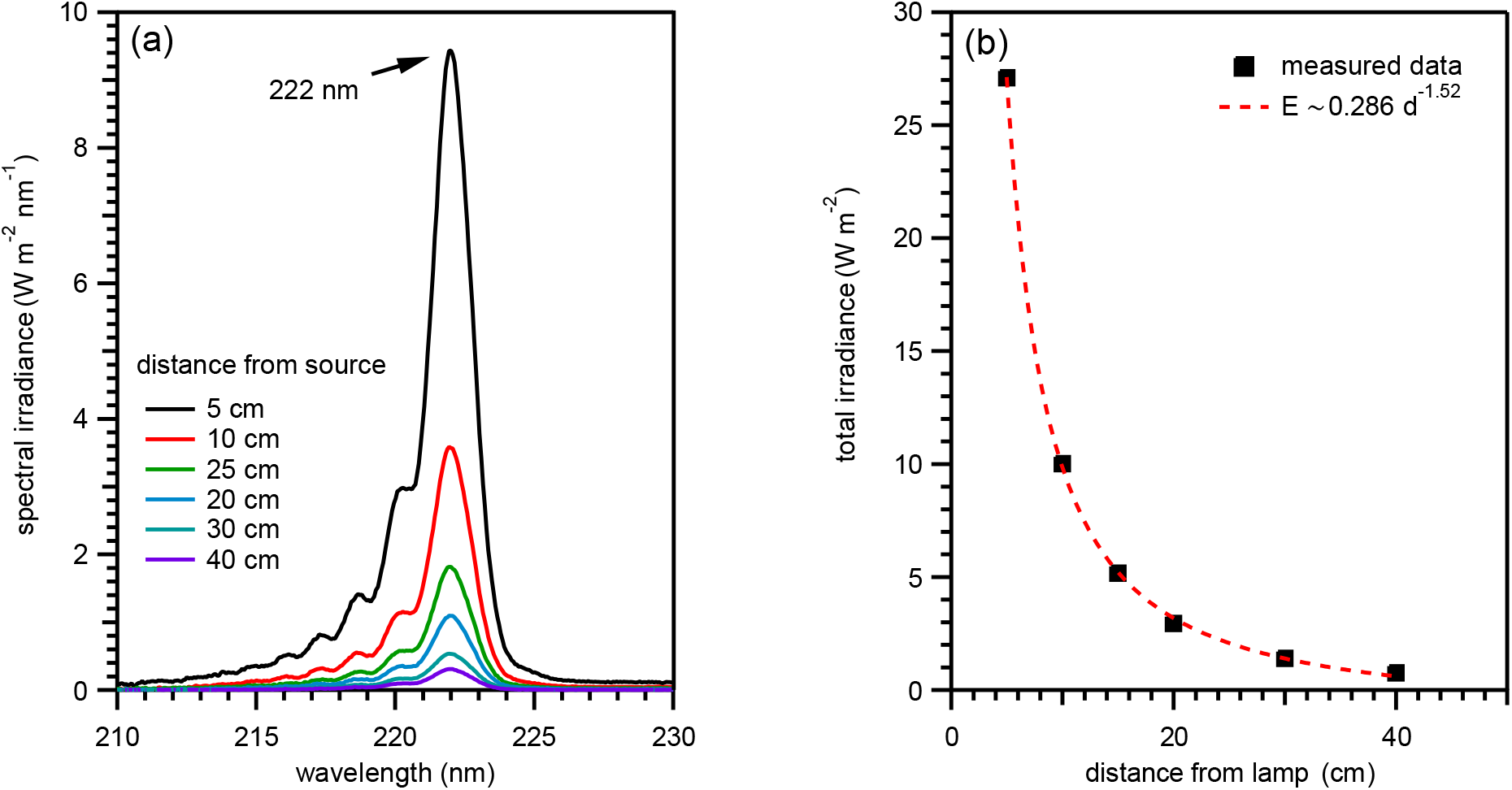
(a) GUV222 lamp emission spectra showing peak emission at 222 nm measured at six different distances. (b) The total irradiance versus distance showing a drop-off proportional to 1/d^1.52^.

The main emission peak is at 222 nm, as reported by other studies examining emission spectra of KrCl lamps^20^, accompanied by a lower-wavelength tail distribution.^21^ By integrating under the spectral irradiance curve over the entire emission range, the total irradiance can be calculated and plotted as a function of distance from the lamp (Figure 1b). The total irradiance in the immediate vicinity of the lamp is high (105 W m^-2^ at 0 cm and 27 W m^-2^ at 5 cm) but drops very quickly with distance. This drop-off follows the relationship, *E* ∼ 1/*d*^1.52^, where *E* is irradiance and *d* the distance from the lamp.

### Measurement and Modeling of O_3_ Production from the GUV222 Lamp

We measured elevated levels of O_3_ in our chamber after four hours of GUV222 lamp operation as shown in Figure 2.

**Figure 2.**
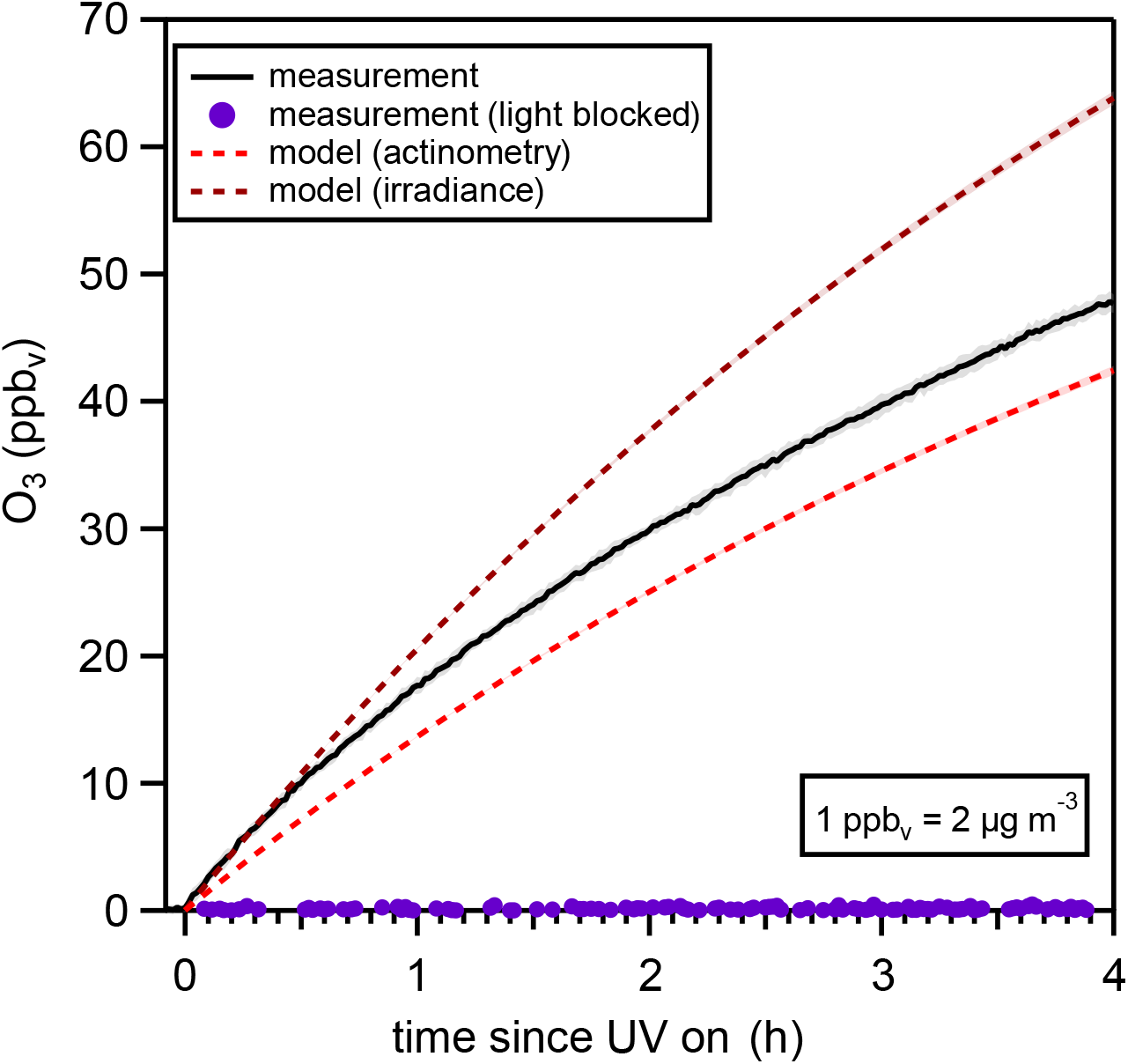
The average O_3_ mixing ratio from seven GUV222 lamp experiments is shown as the solid black line with the variability (2σ) shown by the gray shaded area. The average and standard deviation of seven modeled O_3_ mixing ratios is shown as determined by the irradiance method in dark red and actinometry method in light red. The O_3_ measured from the experiment where the light was blocked is shown in purple.

Four hours after turning the GUV222 lamp on, we observed 48 ppb_v_ ± 1 ppb_v_ (94 μg m^-3^ ± 2 μg m^-3^) of O_3_ in the chamber. To rule out the influence of other physical phenomena related to operation of the GUV222 lamp (e.g., electrical arcing^13^) being responsible for O_3_ production we operated the lamp, for one experiment, with the output of the lamp covered to prevent light from illuminating the chamber. No O_3_ generation was observed in that experiment (Figure 2, purple trace) providing evidence that photolysis of O_2_ at 222 nm was responsible for production of O_3_.

At the end of each experiment the lamp was turned off and the decay of O_3_ was measured (Figure S6). We assume that O_3_ is lost to stainless steel chamber surfaces and homogeneous gas-phase reactions via a first order process. Additionally, some O_3_ is lost via air change which was quantified from SF_6_ decay measurements (≈ 6 %; Figure S8). We determine the rate constant from a linear fit of the natural log of O_3_ mixing ratio versus time (equation 3).

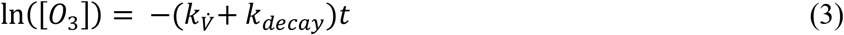

In equation 3, k_decay_ is the first order rate constant for loss of O_3_ to the chamber surfaces and homogeneous gas-phase reaction, 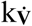 is the air change rate (h^-1^), and t is time. Rates of O_3_ decay (k_decay_) remained relatively constant throughout the experiments only varying by 2 %.

We calculate theoretical O_3_ production from the GUV222 using chemical production and loss and physical loss terms in equation 4.

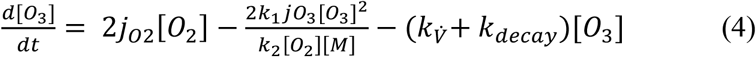

The first term on the right hand side of the equation is the O_3_ production from photolysis of O_2_ at 222 nm, the second term accounts for loss of ozone through the odd-oxygen (O_x_ = O_3_ + O) steady-state (k_1_ = 7.96 × 10^−15^ cm^3^ molecule^-1^ s^-1^; k_2_ = 6.10 × 10^−34^ cm^6^ molecule^-2^ s^-1^), and depositional loss to chamber walls and homogeneous gas-phase reactions is accounted for in the measured k_decay_. The odd-oxygen steady-state is established from the rapid production of oxygen atoms (O) from both O_2_ and O_3_ photolysis (jO_3_ is the photolysis rate constant for O_3_) and the recombining of O with O_2_ to form O_3_.

As shown in equation 4, the photolysis rate of O_2_ drives O_3_ production from the GUV222 lamp and the first-order photolysis rate constant (jO_2_) is strongly dependent on the photon flux (F; equation 5) from the lamp.

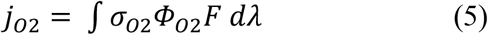

Using the measured irradiance spectrum (Figure 1) from the lamp we calculate an effective O_2_ absorption cross section (σ_O2_)^9^ of 4.30 × 10^−24^ cm^2^ across a wavelength (λ) range between 210 nm and 230 nm (compared to 4.09 × 10^−24^ cm^2^ at 222 nm). The photolysis quantum yield of O_2_ (*Φ*_*O*2_) between 210 nm and 230 nm is unity.^22^ We estimate an effective photon flux (F) from the GUV222 lamp from two different methods: (1) by determining the average of the measured irradiance projected into a cone (irradiance method) and (2) following the method of Peng, et al. (2023)^19^, using chemical actinometry^23^ with tetrachloroethylene (C_2_Cl_4_) as the actinometer (actinometry method).

Briefly, for the irradiance method, we generated an irradiance field within a 31.5 m^3^ cone by expanding the GUV222 lamp irradiance point source axially following the relationship, *E* ∼ 1/*d*^1.52^, and angularly following a relatively tight half-angle of ≈ 55° (equation S4). We then averaged the projected irradiance over the emission volume to get the effective photon flux. For the actinometry method, C_2_Cl_4_ was introduced to the chamber and the GUV222 lamp was turned on for four hours to measure the C_2_Cl_4_ photolysis rate. Using the measured C_2_Cl_4_ photolysis rate, effective cross section (σ_C2Cl4_), and reported photolysis quantum yield (Φ _C2Cl4_), we determined the effective photon flux (equation S8). Between 210 nm and 230 nm, effective GUV222 lamp powers of 32.7 mW m^-2^ and 21.7 mW m^-2^ were determined from the irradiance method and actinometry, respectively. Details of the effective photon flux determination methods are discussed in the supplemental information.

The models both show rapid production of O_3_ early in the experiment and on the approach to steady-state conditions. This rapid rise in O_3_ concentration is due to photolytic production and the relative lack of non-photolytic loss and is consistent with the measured data. For the irradiance method, O_3_ levels are over-predicted by ≈ 33 %. We expect over-estimation of the effective photon flux using this irradiance method because we are not accounting for attenuation of the incident radiation by interactions with the chamber walls. Ma, et al. (2023) recently demonstrated that different types of stainless steel reflect 222 nm light reflect with an efficiency of ≈ 20 %.^24^ The 31.5 m^3^ modeled conical irradiance field slightly extends beyond the chamber walls, but the lamp was positioned in a corner of the chamber such that a large volume of the chamber air was irradiated by the UV light, therefore, our model should be mostly valid. In reality, a majority of the ozone is created within 2 m of the lamp (Table S1), so the cone extending beyond the chamber walls results in small overestimation of ozone production. Accurately accounting for reflectance and exact chamber dimensions would decrease the effective photon flux and thus modeled O_3_ production.

In contrast, for the actinometry method, the model underpredicts O_3_ levels by ≈ 11 %. An effective lamp power of 23 mW m^-2^ (k_decay_= 0.17 h^-1^) would be needed to reconcile the 11% deficit in modeled O_3_ production, which is captured by the measured variability of the effective photon flux determined from actinometry (21.7 ± 1.7 W m^-2^). Despite some discrepancies between modeled and measured O_3_, our calculations provide evidence to suggest the mechanism of O_3_ production from GUV222 lamps is likely photolysis of O_2_ from 222 nm light, and not from other physical phenomena.

### Determination of O_3_ Generation Rates from GUV222 Lamps

In the chamber experiments O_3_ was generated from the GUV222 lamp while simultaneously being lost through air change, gas-phase reactions, and deposition to surfaces. Thus, the O_3_ production rate from the lamp can be determined by solving for the generation rate (GR) in the transient solution to the mass balance equation presented in equation 5.

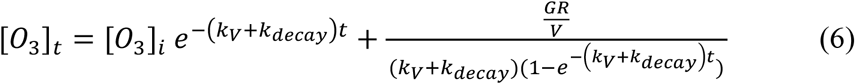

Where [O_3_]_i_ and [O_3_]_t_ are the initial and time t O_3_ mixing ratios, V is the volume of the chamber (31.5 m^3^), and GR is the O_3_ generation rate (μg m^-3^).

Calculated O_3_ production rates from the GUV222 lamp, presented in Table 1, varied within 2%.

**Table 1.** Summary of O_3_ decay constants, air change rates, and O_3_ generation rates.

| Experiment | $k_{\text{decay}} (\text{h}^{-1})$ | $k_V (\text{h}^{-1})$ | GR, $O_3$ generation rate ($\mu\text{g h}^{-1}$) |
| --- | --- | --- | --- |
| 1 | 0.172 | 0.010 | 1126 |
| 2 | 0.176 | 0.014 | 1101 |
| 3 | 0.169 | 0.010 | 1087 |
| 4 | 0.168 | 0.011 | 1087 |
| 5 | 0.167 | 0.012 | 1084 |
| 6 | 0.167 | 0.014 | 1084 |
| 7 | 0.171 | 0.012 | 1104 |
| Average ($\pm 2\sigma$) | $0.170 \pm 0.003$ | $0.012 \pm 0.002$ | $1096 \pm 15$ |

From an average of seven experiments, the O_3_ generation rate from the 222 nm lamp was measured to be 1096 μg h^-1^ ± 15 μg h^-1^.

## Conclusions

We measured the spectral irradiance of a commercial GUV222 lamp from 210 nm to 230 nm showing a peak emission at 222 nm. Results from seven replicate experiments of the single 222 nm commercial GUV222 lamp used in this study yielded a mean O_3_ generation rate of 1096 μg h^-1^. O_3_ generation rates determined in this study could be used to predict O_3_ production and accumulation in indoor spaces from commercial GUV222 lamps like the one used in this study. The results observed in this study apply to this lamp and may vary between unit, manufacturer, and test conditions. For instance, a recent study^24^ measured an O_3_ generation rate from an unfiltered GUV222 lamp nearly ten times lower than the average value reported in this study however that study did not account for the dynamic deposition of O_3_ to chamber walls which likely resulted in lower measured O_3_ generation rates. Like the losses of O_3_ to chamber walls and gas-phase reactions observed in this study, similar reactive losses of O_3_ generated from GUV222 devices would be expected in real indoor environments with potential impacts for by-product formation that would affect indoor air quality.^25^ Based on the results from this study we suggest more measurements of O_3_ production should be made from commercial air cleaning devices employing GUV222 lamps in both real indoor and laboratory settings.

## Data Availability

All data produced in the present study are available upon reasonable request to the authors.
All data produced in the present work are contained in the manuscript.

## Disclaimer

Certain equipment, instruments, or materials, commercial or non-commercial, are identified in this paper in to specify the experimental procedure adequately. Such identification is not intended to imply recommendation or endorsement of any product or service by NIST, nor is it intended to imply that the materials or equipment identified are necessarily the best available for the purpose.

## Acknowledgments

We would like to acknowledge James Norris and Peter Trask for calibration of the ozone instrument used in this study. We would like to thank Howard Yoon and Cameron Miller for assistance with irradiance calibrations of our UV spectroradiometers. We thank and acknowledge Jose Jimenez for providing recommendations for experimental design.

