## supplement for "Ozone Generation from a Germicidal Ultraviolet Lamp with Peak Emission at 222 nm"

**Supplemental Information for: Ozone Generation from a Germicidal Ultraviolet Lamp with Peak Emission at 222 nm**

**Determination of the effective photon flux from the GUV222 lamp**

Ozone (O_3_) production from 222 nm light is driven by oxygen photolysis.^1^ Equation S1 shows the rate equation for the rate of oxgen loss from photolysis at 222 nm and equation S2 shows the equation determining the first-order rate constant for oxygen photolsyis (jO_2_).

$\frac{d[O_{2}]}{dt}= -j_{O2}[O_{2}]$	(S1)

$j_{O2}= \int\sigma_{O2}\Phi_{O2}F d\lambda$	(S2)

In order to calculate j_O2_ we determined an effective oxygen absorption cross section ($\sigma_{O2}$) by taking an irradiance intensity weighted average of the $\sigma_{O2}$ between the wavelengths of 210 nm to 230 nm. The oxgen photolysis quantum yield ($\Phi_{O2}$) is equal to unity in this region.^2^ Direct measurement of the photon flux, F, from the lamp throughout the chamber is difficult, so we used two methods to estimate it: (1) projection of the directly measured spectral irradiance into an idealized cone and (2) chemical actinometry using tetrachloroethylene (C_2_Cl_4_) as the actinometer.

**Averaging of the irradiance field projected into a cone:**

The total irradiance in the immediate vicinity of the lamp is high (105 W m^-2^ at 0 cm and 27 W m^-2^ at 5 cm) but drops very quickly with distance. For distances of at least 5 cm, this drop-off follows the relationship,

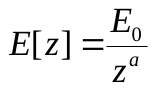
 (S3)

where *E*[*z*] is the total irradiance as a function of normal distance from the lamp, *z*. From the fit in shown in Fig. 1b, we find
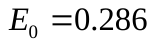
W/m^(2-a)^ and *a* = 1.52. To approximate the angular distribution of the lamp emission, irradiance measurements were performed at several distances away from the lamp while the lamp was rotated away from normal in 5° increments on a rotation stage. S1 shows the relative irradiance for a given projection angle, θ, normalized to the irradiance at the same distance from the lamp at θ = 0°, E_max_, (i.e., lamp facing the detector). This plot includes all measurements at distances of 10 cm, 15 cm, 20 cm, 30 cm, and 40 cm.

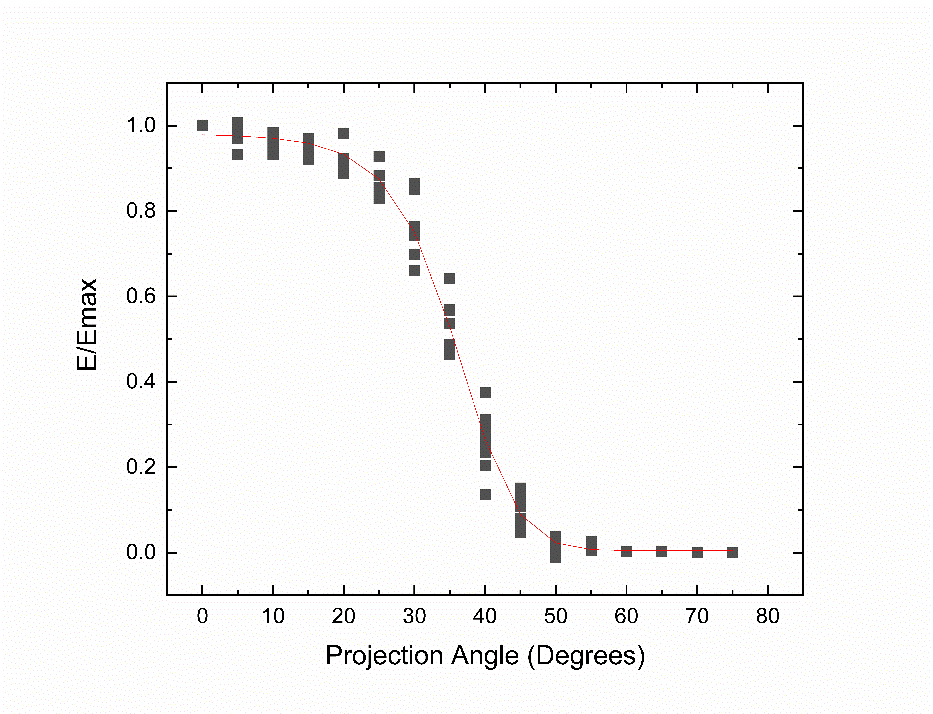

**Figure S1.** Relative irradiance from GUV222 lamp at various projection angles from 0 degrees to 75 degrees. The irradiance becomes negligible at 55 degrees.

The trend indicates that the irradiance rapidly drops off with θ, independent of the distance from the lamp; therefore, most of the UV photons are within a relatively tight half-angle of ≈ 55°. To approximate this trend with a mathematical function, this data was fit to a sigmoidal curve of the form:

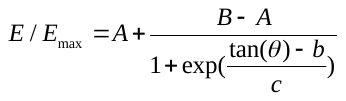
 (S4)

with the fit parameters given by: *A* = 0.00479, *B* = 0.9809, *b* = 0.7180, *c* = 0.1196. Equations S3 and S4 can be combined to produce the approximate irradiance distribution of most of the UV emitted photons into a cone in space directly in front of the lamp. To do this, we define this 3D spatial irradiance as
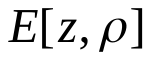
, where *ρ* is the radius of the circle the lamp projects at distance *z*:

$E\left[ z,\rho\right]= \left\{ \begin{aligned} \frac{E_{0}}{\left( \sqrt{z^{s}+\rho^{2}} \right)^{a}}\cdot\left( A+\frac{B-A}{1+{exp}^{\left( \frac{\frac{\rho}{z}-b}{c} \right)}} \right) z>z_{min} \\ 105 W / m^{2} z<z_{min} \end{aligned} \right.$	(S5)

Here, we place a lower bound on z, z_min_, because the highest irradiance is at the lamp surface to prevent this function from producing ever higher E values as z approaches 0.

With
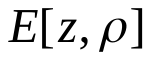
 defined, we can now calculate an average irradiance value in space as follows:

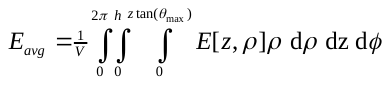
 (S6)

*V* is the total volume of the emission cone such that *V* = 31.5 m^3^, the volume of our test chamber, *h* is the maximum height of the cone for this volume, *h* = 2.45 m, and *θ*_max_ = 55° as determined by the angular measurements. Since all the UV-generated ozone is being forced to circulate and fill the entire test chamber volume, calculating an average irradiance over this volume is a reasonable assumption. In this calculation, we are also ignoring reflections off the chamber walls and ignoring that the 31.5 m^3^ modeled cone extends beyond the chamber walls. However, the lamp was positioned in a corner of the chamber such that the majority of the light is projected into the volume of the chamber and therefore we believe the errors due to side-wall reflections are not large. We solved Eq. S6 numerically and obtained a volume averaged irradiance of

E_avg_ = 0.0327 W m^-2^, corresponding to an average photon flux φ = 3.65 × 10^16^ photons m^-2^ s^-1^.

**Table S1.** Modeled O_3_ production as a function of z-axis distance from the lamp within the conical irradiance field.

| z (m) | E_avg_ (W m^-2^) | O_3_ concentration in lamp volume (ppb_v_) | O_3_ concentration for chamber (ppb_v_) | Percent of O_3_ concentration compared to z = 2.41 m |
| --- | --- | --- | --- | --- |
| 0.5 | 0.3670 | 615.9 | 5.48 | 9.74 |
| 1.0 | 0.1280 | 214.7 | 15.28 | 27.1 |
| 1.5 | 0.0690 | 115.9 | 27.90 | 49.6 |
| 2.0 | 0.0446 | 74.85 | 42.60 | 75.7 |
| 2.25 | 0.0373 | 62.58 | 50.75 | 90.1 |
| 2.33 | 0.0354 | 59.35 | 53.45 | 95.0 |

**Chemical actinometry**

Chemical actinometry has an extensive history of use in laboratory studies of chemical kinetics to measure the effective photon flux capable of initiating a chemical reaction^3^ or for determining photolysis quantum yields.^4^ Chosing an actinometer is dependent on different experimental considerations, but principly a chemical actinometer should absorb strongly in the wavelength region of interest. Figure S1 shows relatively high levels for the tetrachloroethylene (C_2_Cl_4_) absorption cross section, $\sigma_{C2\mathrm{Cl}4}$, in our region of interest so we chose C_2_Cl_4_ as a chemical actinometer for indirect measurement of the photon flux from the GUV222 lamp.

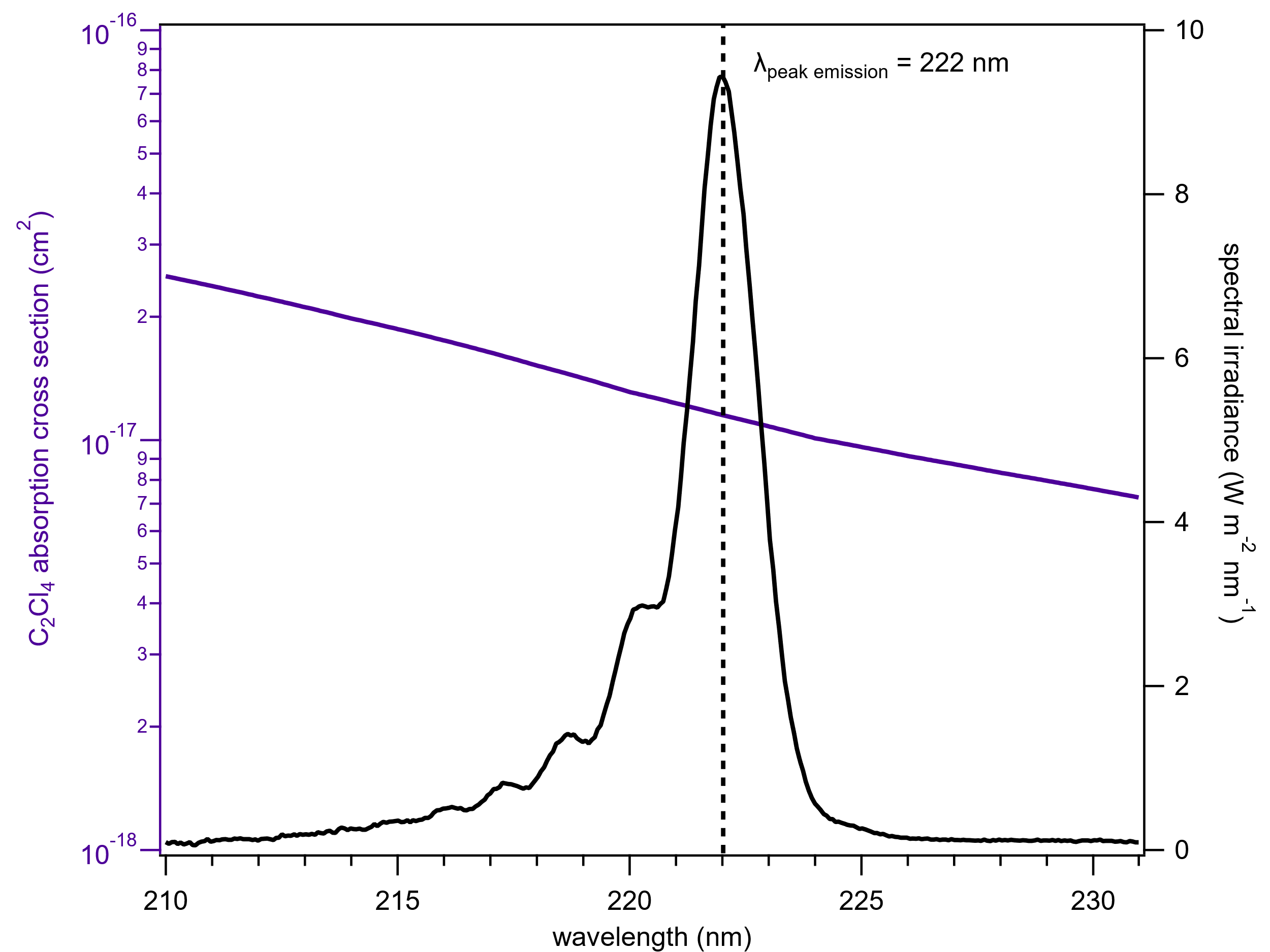

**Figure S2.** C_2_Cl_4_ asborption cross section ($\sigma_{C2\mathrm{Cl}4}$) versus wavelength. The dotted line shows the absorption cross section at 222 nm^5^. After multiplying $\sigma_{C2\mathrm{Cl}4}$ by the normalized spectral irradiance, we calculate an effective $\sigma_{C2\mathrm{Cl}4}$of 1.24 x 10^-17^ cm^2^ for the GUV222 lamp.

Additionally, the photolysis quantum yield of C_2_Cl_4_ at 222 nm is unity. We measured the rate of C_2_Cl_4_ photolysis in the chamber from GUV222 light and, using equation S2 (subsituting values for C_2_Cl_4_ in place of O_2_), determined the effective photon flux. Though the actinometer used in this study is different, determining the effective photon flux using actinometry is a method we adopted from Peng, et al. (2023)^6^.

Liquid C_2_Cl_4_ was injected into a heated stream of ultra zero air and supplied to the 31.5 m^3^ chamber to producing levels of C_2_Cl_4_^+^ that were measured by proton-transfer mass spectrometry (Figure S3) for four of the experiments described in the main text.

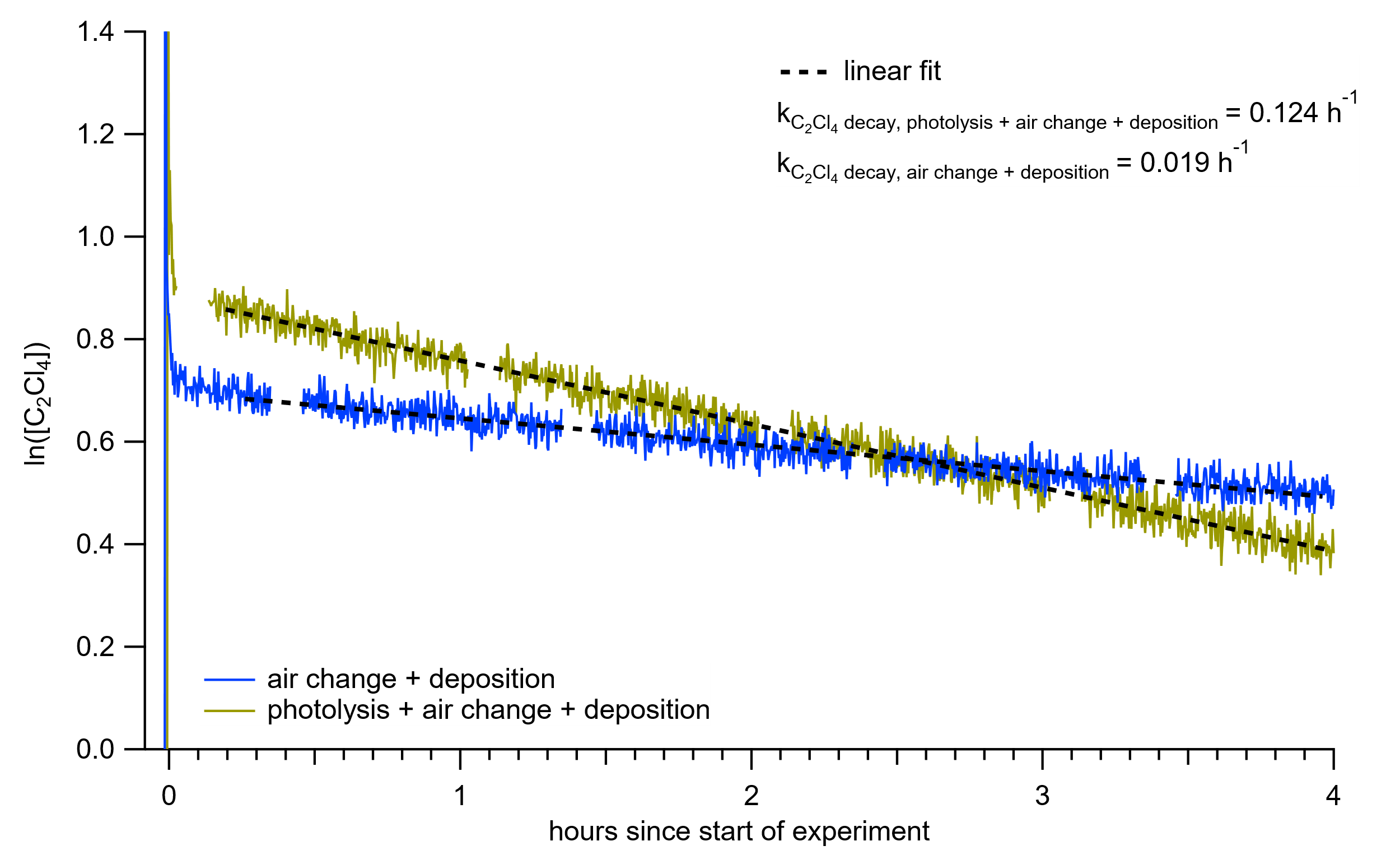

**Figure S3.** C_2_Cl_4_^+^ signal produced from introduction of C_2_Cl_4_ to chamber. The first four hours show decay associated with both photolysis and loss to chamber walls or air change (yellow). After the GUV222 lamp has turned off the rate of loss to the chamber walls and air change can be observed (k_loss_). About 15 % of the loss of C_2_Cl_4_ when the GUV222 lamp was on was from the combined effects of deposition to chamber walls and air change.

The rate of C_2_Cl_4_ photolysis was determined from equation S7:

$\ln\left( \left[ C_{2}Cl_{4} \right] \right)= -\left( k_{photolysis + deposition + air change}- k_{deposition+ air change} \right)t$	(S7)

The difference between the two k values in equation S7 gives the first-order photolysis rate of tetrachloroethylene (jC_2_Cl_4_). The effective photon flux was then determined from equation S8:

$F= \frac{jC2Cl4}{\sigma_{C2Cl4}}$	(S8)

The effective photon flux determined via actinometry was 2.43 x 10^12^ photons cm^-2^ s^-1^ (GUV222 equivalent lamp power of 21.7 mW m^-2^).

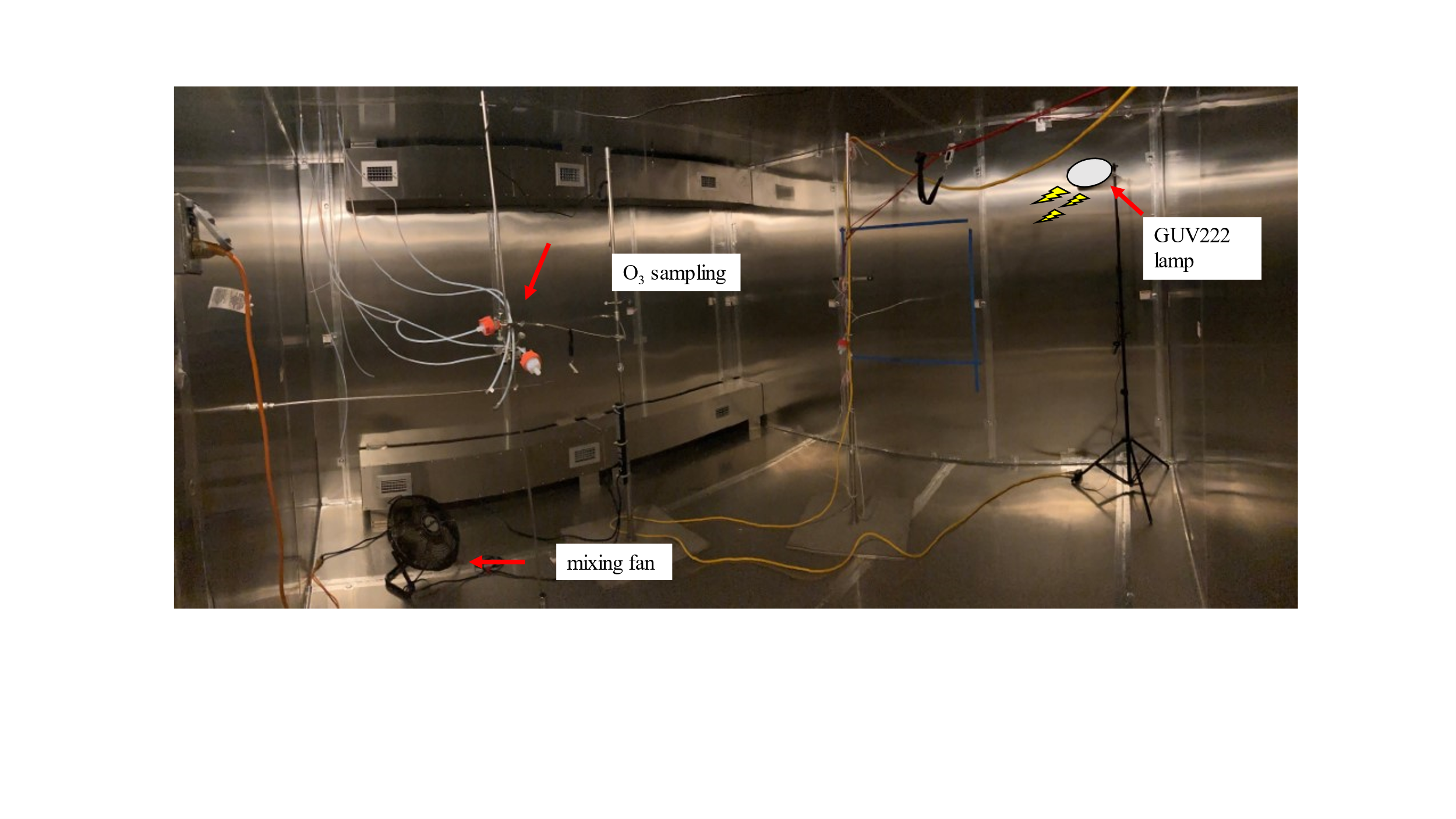

**Figure S4.** Picture of the 31.5 m^3^ stainless steel environmentally controlled walk-in chamber with the GUV222 lamp set up in the corner. The lamp was pointed to the center of the room at a downward angle of ≈ 20°.

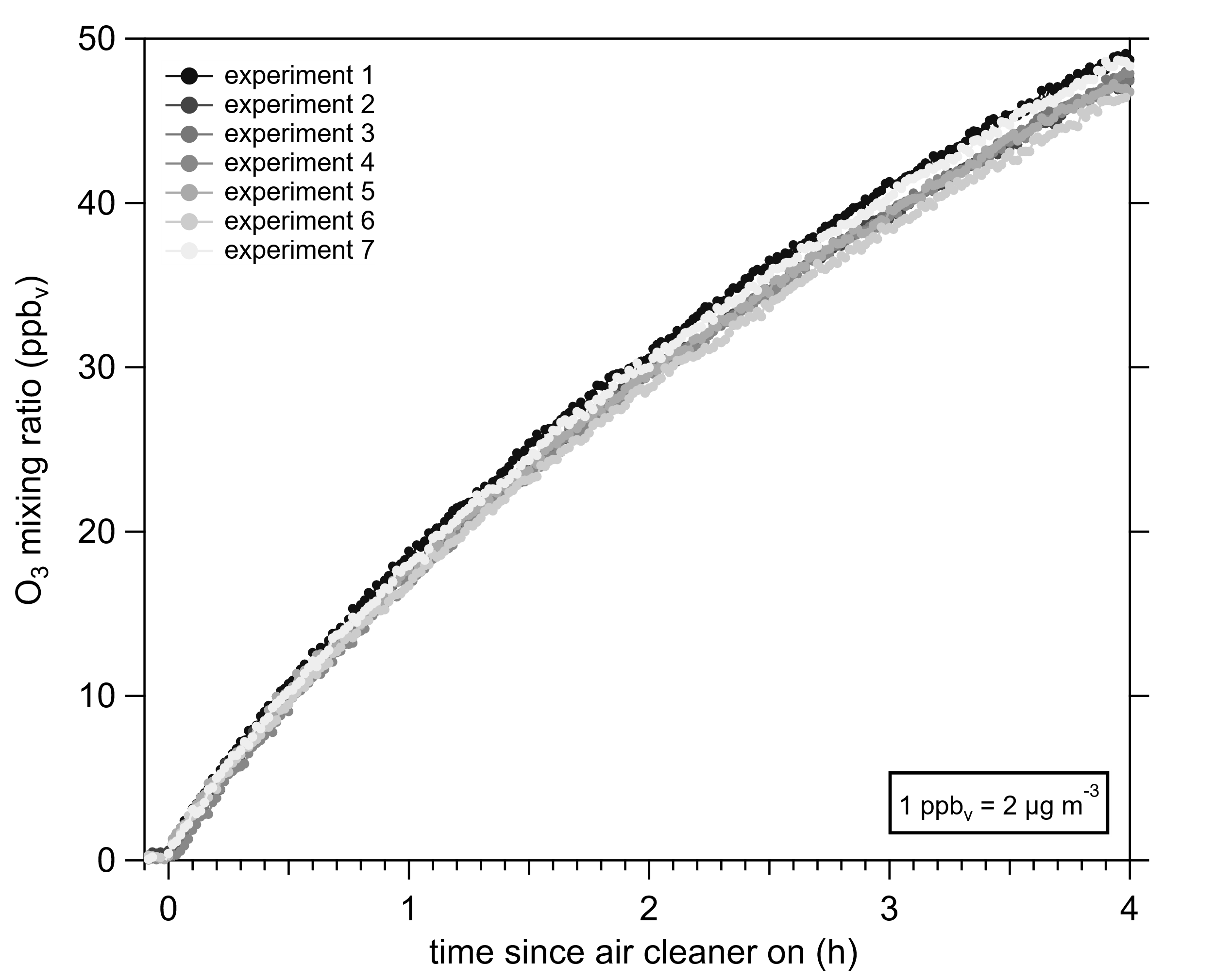

**Figure S5.** O_3_ generation observed from each experiment with the GUV222 lamp.

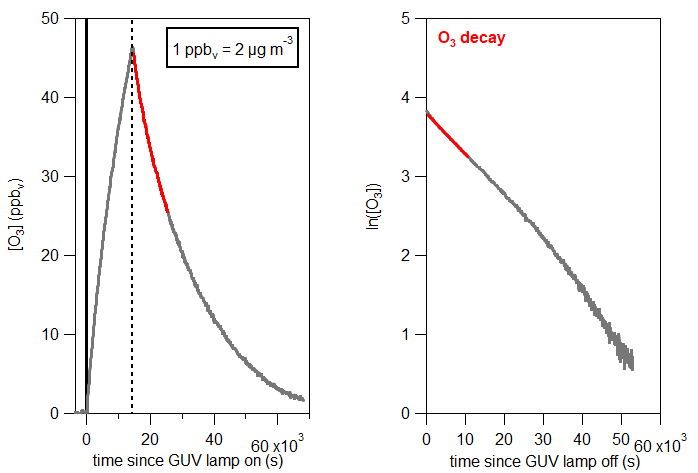

**Figure S6.** Example of the measured O_3_ mixing from experiment 7 (left panel). The time when the lamp was turned off is indicated by the vertical dotted line. First-order rate constants for O_3_ decay are determined from a linear fit to the first three hours (red traces) of the natural log of the O_3_ mixing ratio after the lamp was turned off.

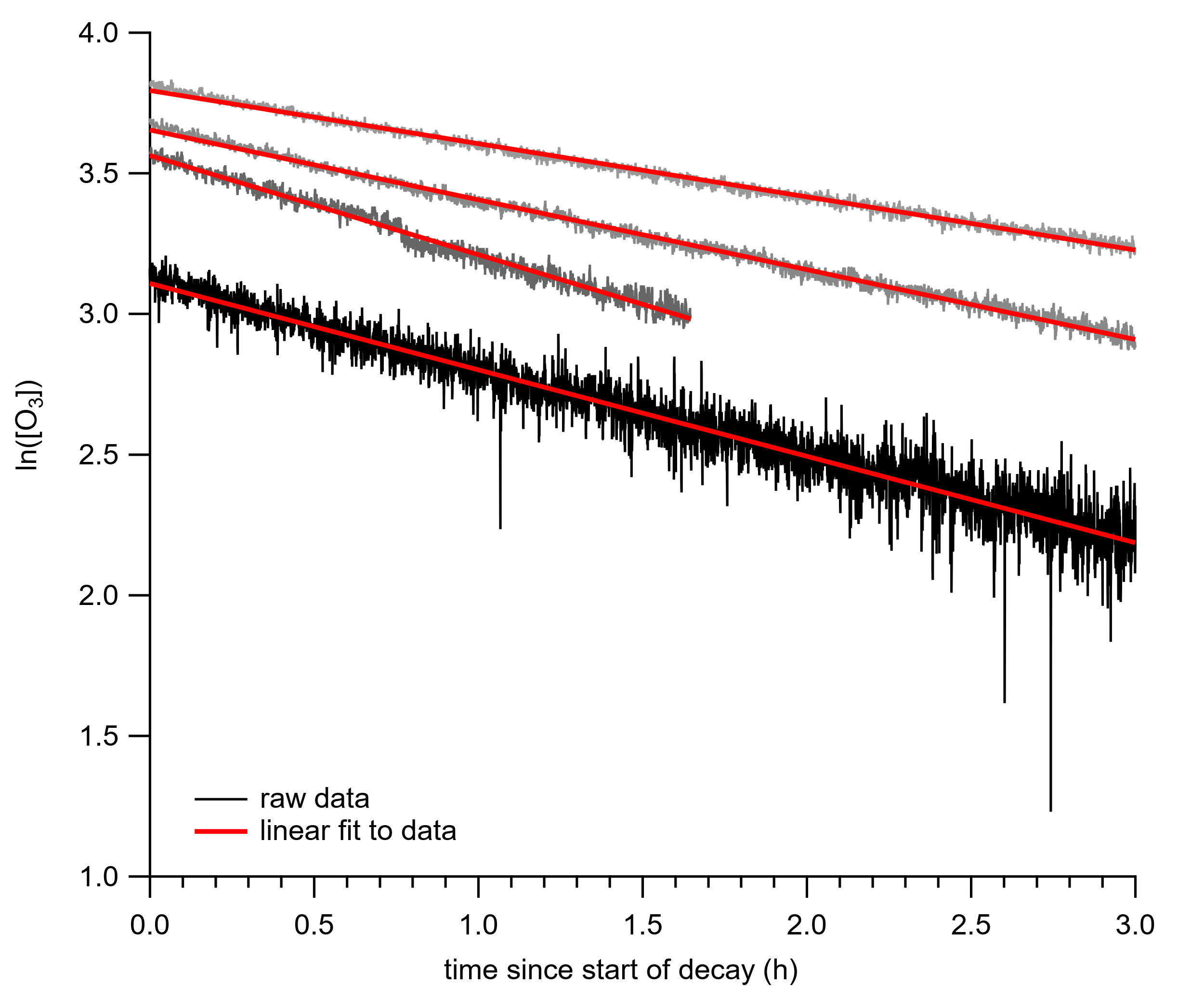

**Figure S7.** Linear fitting of the natural log of the O_3_ mixing ratio versus time since the air cleaner (O_3_ source) was turned off. The first-order rate constant determined from this fitting describes the combined homogenous gas-phase and heterogeneous gas-to-surface losses as well as loss via air change and when multiplied by the ratio of the chamber volume to area gives the O_3_ deposition velocity (cm s^-1^).

**Chamber ventilation and air change rate (V̇) measurement**

Prior to each experiment outdoor air was recirculated through the chamber to control the temperature and humidity. We operated the chamber to achieve a temperature of 20 °C and 50 % relative humidity. At the beginning of each experiment temperature and humidity control was stopped and the vents controlling the recirculation of air were closed. During the experiments air change occurred through leakage points both in the chamber and in the duct work to the chamber.

The air change rate ($\dot{A}$) of the chamber was determined in four experiments using the tracer decay method (see ASTM E741)^7^ via injection of 2 L of 10 % sulfur hexafluoride (SF_6_) in a nitrogen balance at the beginning of the experiment. SF_6_ was measured by hydronium ion (H_3_O^+^) proton-transfer time-of-flight mass spectrometry (PTR-MS). To the best of our knowledge SF_6_ measurements by PTR-MS have never been reported. We measure SF_6_ product ions SF_3_O^+^, SF_3_^+^, SF_2_^+^, and SF^+^ in order of decreasing sensitivity. We measure SF_6_ via SF_3_O^+^ (m/z 104.962) with a sensitivity ≈ 8 cps ppb_v_^-1^ (LOD = 350 ppt_v_; S/N = 3 and 10 s measurement resolution). An example of the determination of the first-order constant for the air change rate (k**_V̇_**) is shown in Figure S8 from the measurment of SF_3_O^+^ (from SF_6_) decay by PTR-MS.

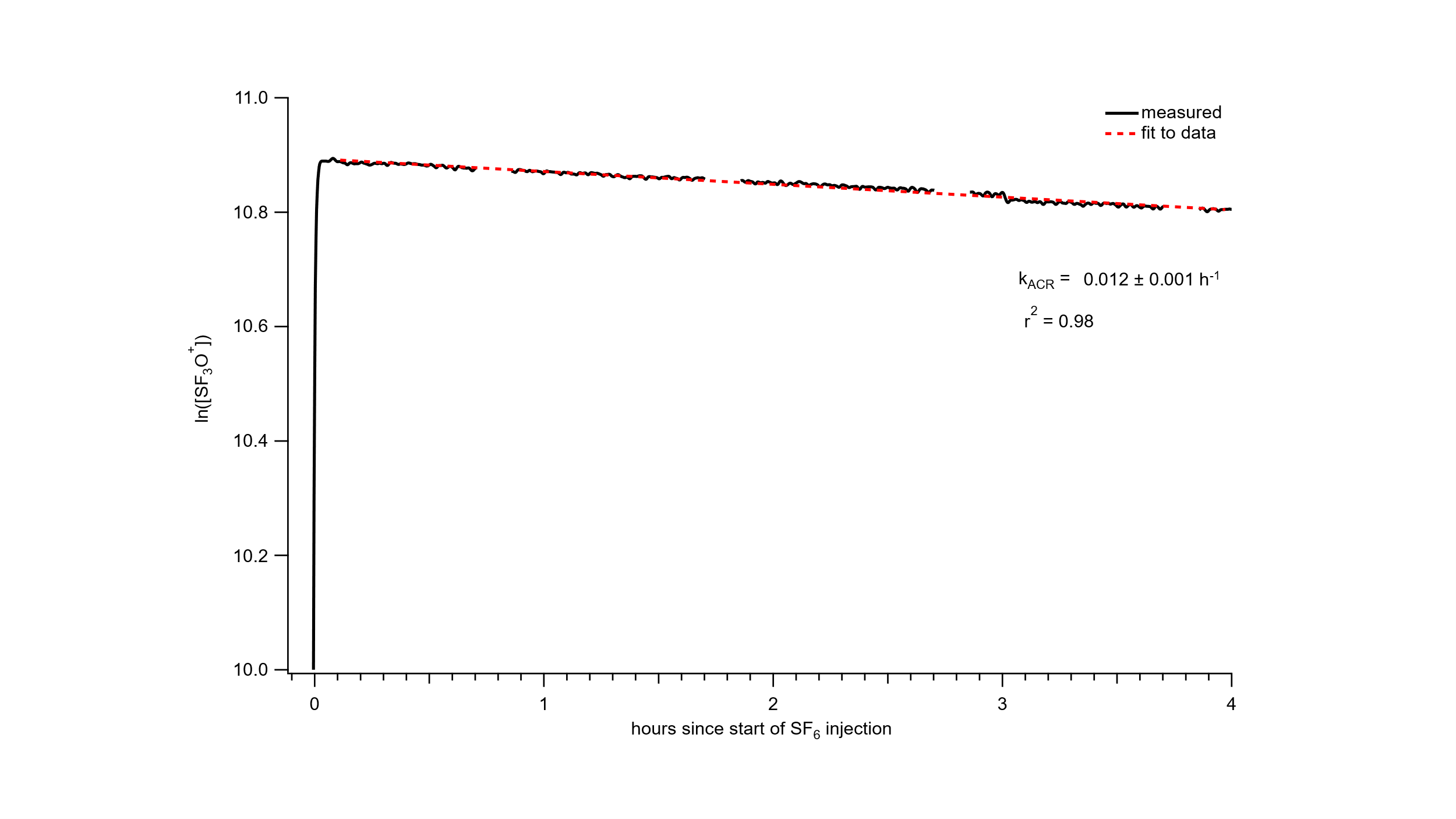

**Figure S8.** The natural log of the signal from SF_3_O^+^, produced from PTR-MS ionization of SF_6_, decays as a first order loss. We determine V̇ as 0.012 h^-1^ ± 0.001 h^-1^ from this injection of SF_6_ into the chamber.
